# Axonal Degeneration Across the Alzheimer’s Disease Spectrum: A Longitudinal MRI and Fluid Biomarker Study

**DOI:** 10.1101/2025.08.13.25333630

**Authors:** Zhaoyuan Gong, John P. Laporte, Alexander Y. Guo, Jonghyun Bae, Noam Y. Fox, Angelique de Rouen, Nathan Zhang, Aaliya Taranath, Rafael de Cabo, Josephine M. Egan, Luigi Ferrucci, Mustapha Bouhrara, the Alzheimer’s Disease Neuroimaging Initiative

## Abstract

With global dementia rates rising sharply, there is an urgent need for sensitive biomarkers to detect cognitive changes and predict dementia risk. White matter degeneration, especially axonal loss, is increasingly recognized as an early hallmark of Alzheimer’s disease (AD), but its temporal trajectory and its relationship with cognition have not been established. We utilized a novel MRI-derived Axonal Density Index (ADI) to longitudinally investigate axonal degeneration and cognitive decline in the ADNI cohort. Linear mixed-effects models showed cognitively impaired individuals had lower baseline ADI and faster axonal degeneration compared to cognitively normal subjects. In cognitively impaired individuals, higher baseline ADI predicted slower prospective cognitive deterioration and lower dementia risk, while greater longitudinal ADI declines correlated with cognitive worsening and increased dementia risk. Notably, ADI outperformed cerebrospinal fluid biomarkers of AD pathology in predicting cognitive outcomes. Our original findings position axonal degeneration as an early AD feature and ADI as a promising biomarker for early detection, disease phenotyping and monitoring, and intervention targets.

## Introduction

Anti-amyloid interventions have failed so far to deliver tangible and substantial therapeutic benefits, highlighting the need for a more comprehensive understanding of Alzheimer’s disease (AD) mechanisms (1–3) that extends beyond the traditional amyloid (A), tau (T), neurodegeneration (N) (AT(N)) framework (4, 5). This also emphasizes the importance of identifying reliable biomarkers that can detect AD in its prodromal phase and assessing the risk of progression at an early stage before diagnostic symptoms are developed. Accurate and quantitative biomarkers at this early stage are also essential for monitoring disease progression and optimizing therapeutic targets that could potentially slow AD progression or prevent its onset in at-risk individuals. The classical AT(N) biomarker paradigm defines AD based on the development of amyloid-β (Aβ) plaques, neurofibrillary tau tangles, and structural atrophy in brain regions critical for memory and cognition (6). These hallmark pathological features are considered to represent distinct stages of the disease process, with Aβ plaques typically appearing first, followed by tau tangles and, eventually, structural atrophy. While these biomarkers can readily be assessed using positron emission tomography (PET) imaging or cerebrospinal fluid (CSF) analysis, these methods are invasive, expensive and not widely accessible, making them less ideal for routine screening or large-scale population studies. Furthermore, macrostructural changes, such as hippocampal and cortical atrophy, are likely to manifest at more advanced stages of the disease, making them less reliable for early detection and intervention. Importantly, the current AT(N) framework overlooks the complexity of AD as reflected by the involvement of multiple biological pathways beyond amyloid and tau, including neuroinflammation, mitochondrial dysfunction, demyelination, axonal degeneration, synaptic loss and vascular changes, all of which contribute to the progressive cognitive decline in AD, likely with different degree of relevance across individuals (7, 8). These limitations highlight the need for further expansion of the AT(N) framework, preferentially by the development of noninvasive, sensitive, specific and cost-effective biomarkers.

Recent evidence suggests that white matter (WM) alterations appear early in AD and other neurodegenerative disorders, contributing to cognitive, motor, and autonomic impairments (9–11). Indeed, brain autopsy studies found that WM degradation is a common feature of AD (12–17), with single-cell transcriptomics studies revealing perturbed WM-related genes (18), reduced major myelin proteins (19), and higher inflammation (20). Reduced WM integrity was observed in a transgenic mouse model of amyloidosis (21) and experimentally induced myelin damage caused accelerated parenchymal amyloidosis, suggesting a role of WM damage in the development of Aβ. For tau pathology, alterations in WM integrity were observed in a transgenic mouse model of fibrillar tau (22, 23), preceding the emergence of tau pathology (22), with clinical investigations suggesting a role of WM hyperintensities in tau pathology in the AD spectrum (24). Additionally, clinical studies in genetic forms of AD, including autosomal dominant AD and Down syndrome, highlight impairments in WM integrity decades before the onset of symptoms (9, 25, 26). These collective findings suggest that WM alteration is an important component of AD pathophysiology. WM degeneration disrupts neural pathways, impairing the communication between brain regions whose integrity is essential against cognitive decline, mobility, and autonomic dysfunction, such as troubling cardiovascular regulation and bladder control (27–29). Despite accumulated scientific progress, the temporal sequence of cerebral WM microstructural changes in relation to cognition and disease progression remains poorly understood, including in early onset and sporadic AD. Current techniques in characterizing WM tissue integrity focus on mapping myelin sheaths and axons. Recent preclinical and clinical studies shed light on the role of myelin in cerebral aging and AD (27–31) but the relevance of axonal integrity deterioration remains unclear. Investigating axonal degeneration in mild cognitive impairment (MCI) is crucial because it represents one of the earliest clinical manifestations along the AD continuum, potentially preceding overt brain atrophy or significant cognitive decline associated with dementia. Understanding these early microstructural changes may reveal mechanisms driving disease progression, identify novel therapeutic targets aimed at preserving WM integrity, and support the development of noninvasive biomarkers for early diagnosis and risk stratification (32). Moreover, the degree of axonal degeneration may help in disease phenotyping and in identifying subgroups with specific clinical courses, and possibly those that may be more responsive to specific interventions aiming at preserving or restoring WM integrity. This raises a critical question: Are individuals with a clinical diagnosis of cognitive impairment but higher axonal health protected from accelerated cognitive decline and increased dementia risk? Addressing this question requires a reliable and sensitive in vivo measure of axonal integrity.

There is significant interest in leveraging advanced neuroimaging techniques, such as diffusion MRI (dMRI), to noninvasively assess human brain microstructure degeneration, which might precede macrostructural changes and cognitive symptoms by decades (33–35). These methods have the potential to assess WM integrity deterioration with high sensitivity and to provide insight into its role in cognitive decline and neurodegenerative diseases. While various MRI methods can probe cerebral WM microstructural integrity, including relaxation times and diffusion tensor imaging (DTI), only a few methods can specifically quantify axonal density. Axonal density is a quantitative measure that represents the fraction of axonal water relative to the total water content within each brain voxel. It is expected to decline as a result of axonal degeneration and subsequent loss of axonal water. The most clinically adopted techniques are the Neurite Orientation Dispersion and Density Imaging (NODDI) and the Standard Model of Diffusion dMRI (36, 37). Both methods are based on a multicompartment model that distinguishes between water diffusivities within axons and in the extracellular space, providing an estimate of the axonal density index (ADI), a proxy of axonal integrity and health in WM. To address increasing criticisms regarding the physiological reliability of NODDI, we recently introduced a new method, called constrained NODDI (C-NODDI), providing physiologically realistic ADI values in WM that strongly correlate with neurofilament light chain (NfL) concentration level which itself is a plasma biomarker of axonal degeneration (38), with higher NfL values associated with lower ADI. However, unlike widely adopted but nonspecific DTI, these methods require multishell dMRI data, which are now routinely acquired in clinical investigations, including the Alzheimer’s Disease Neuroimaging Initiative (ADNI).

Using the longitudinal ADNI multishell dMRI, CSF and cognitive data and linear mixed-effects regression analyses, we examined the longitudinal changes in ADI as measured using NODDI, C-NODDI or SMI in cognitively impaired (CI) patients, including MCI and AD, and cognitively normal (CN) participants. We also evaluated whether baseline ADI level predicts future changes in cognition and dementia risk, measured by the Mini-Mental State Examination (MMSE) and Clinical Dementia Rating – Sum of Boxes (CDR-SB) scores, respectively. Further, we investigated the association between longitudinal changes in ADI and longitudinal changes in cognition and dementia risk. Finally, we conducted similar analyses using CSF biomarkers of AD pathology and compared results with those obtained by ADI. We hypothesized that: (1) C-NODDI provides a powerful noninvasive imaging biomarker to detect axonal degeneration early in the course of AD and to distinguish ADI trajectories between CN and CI, while also predicting cognitive changes with higher or comparable performance to those derived using biomarkers from CSF or other dMRI techniques. (2) Subjects with a clinical diagnosis of MCI or AD who exhibit higher axonal density will exhibit a less longitudinal decline in cognition and a lower dementia risk.

## Results

### Cohort characteristics

Cohort data availability, longitudinal distribution, MRI biophysical models, and subject characteristics are summarized in Fig. 1. After excluding scans with imaging artifacts or cognitive status changes during longitudinal follow-up, the final cohort included 205 participants and 325 available multishell dMRI measurements (Fig. 1a). Among the participants, 117 were cognitively normal (CN) and 88 were cognitively impaired (CI; including 74 MCI and 14 AD). A total of 82 participants underwent longitudinal assessment, with 50 participants having one follow-up, 25 having two follow-ups, and 6 having three follow-ups from baseline (Fig. 1b). At baseline, age and sex distributions differed significantly between CI and CN participants (p = 0.017 for age; p = 0.008 for sex) (Fig. 1d), with CI participants being older and more likely to be male compared to CN participants. Additionally, males had longer follow-up durations than females (Fig. 1d). Each multishell dMRI scan was processed using SMI, NODDI, and C-NODDI techniques to generate corresponding ADI values (ADI_NODDI_, ADI_C-NODDI_, and ADI_SMI_) in whole-brain white matter (WM), which served as the region of interest in this study (Fig. 1c). The details of image processing pipeline are described in Supplementary Fig. S1.

**Figure 1.**
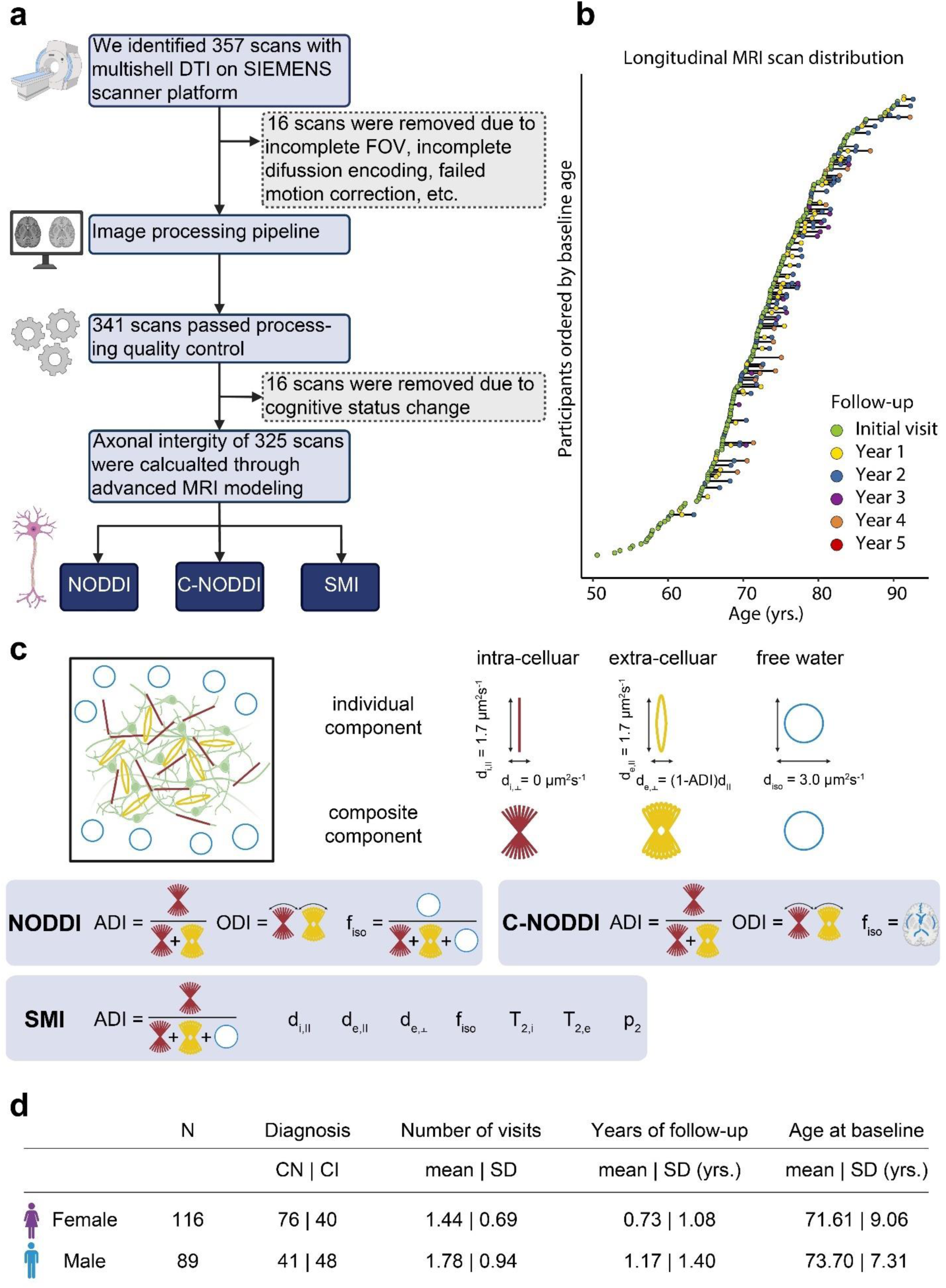
Overview of data acquisition, processing, distribution, diffusion MRI models, and subject characteristics. **a** The flow chart outlines the data acquisition and processing steps. On 01/30/2024, 357 multishell DTI MRI (dMRI) scans on SIEMENS scanners were downloaded. Sixteen scans were excluded after failing quality control and preprocessing steps, and another 16 scans were discarded due to changes in the diagnostic group. The final dataset includes 325 longitudinal scans from 205 subjects over a maximum span of 4.16 years. **b** The longitudinal distribution of dMRI scans is shown, with participants ordered by the age at their first dMRI scan. Time points, from the initial visit to the fifth follow-up, are color-coded. **c** Preprocessed images were input into three different biophysical models to estimate corresponding axonal density index (ADI) maps, a quantitative biomarker of axonal density/integrity. These models are: neurite orientation dispersion and density imaging (NODDI), constrained NODDI (C-NODDI), and Standard Model Imaging (SMI). **d** Table providing summary statistics of the longitudinal dMRI data stratified by sex and cognitive status (CN; cognitively normal, CI; cognitively impaired). Age and diagnosis at baseline were statistically different between males and females. Males had a longer mean follow-up duration, and a slightly higher number of visits as compared to females. However, while the number of CI patients was relatively equivalent between both sexes, the cohort included a higher number of CN females.

### Trajectories of ADI over time in CN and CI

The longitudinal distribution of dMRI data anchored at their baseline scan is shown in Fig. 2a. Representative examples of ADI_NODDI_, ADI_C-NODDI_, and ADI_SMI_ maps from one CN and one CI subject at baseline and at one- and two-year follow-ups are displayed in Fig. 2b. Derived ADI maps from NODDI and C-NODDI showed lower regional values in the CI subject compared to the CN subject. Moreover, the CN subject exhibited minimal regional variation in ADI over time, while the CI subject demonstrated noticeable decreases in regional values. Interestingly, ADI_SMI_ values in the CI subject were slightly higher than in the CN subject and showed minimal change over time in both groups. As expected, ADI_NODDI_ values were substantially higher than those derived from C-NODDI or SMI, with values exceeding 70% in several cerebral WM structures, which are not physiologically plausible as discussed in (38).

**Figure 2.**
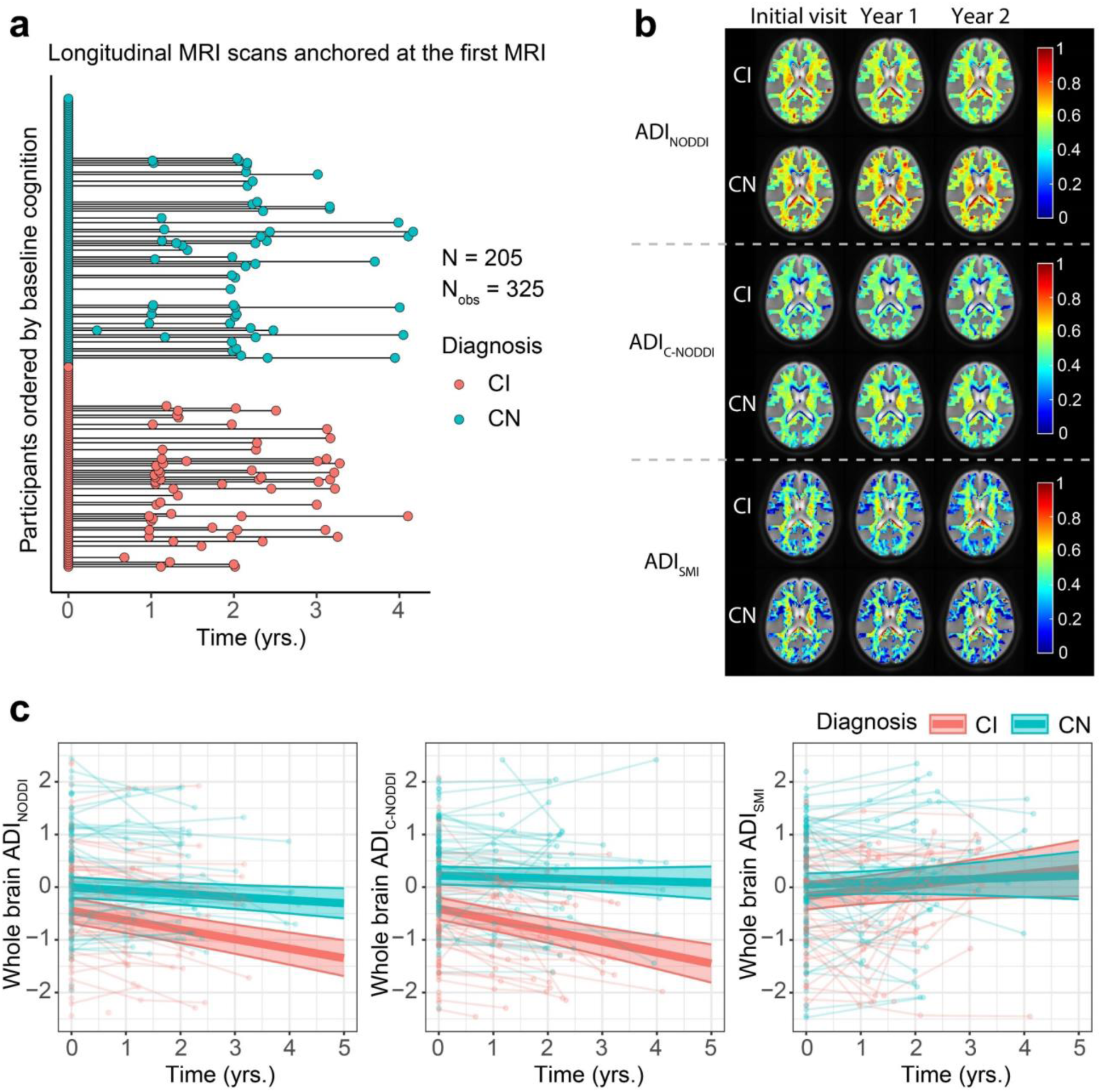
Characterization of longitudinal trajectories of axonal integrity in cognitively normal (CN) and cognitively impaired (CI) subjects. **a** The longitudinal distribution of diffusion MRI (dMRI) scans anchored at the first dMRI scan of each subject, with CN and CI groups color-coded. Both the CN and CI groups have similar longitudinal distributions, with many of the participants having over 3 years of follow-up dMRI measurements from baseline. **b** Representative axonal density index (ADI) maps derived using the neurite orientation dispersion and density imaging (NODDI), constrained NODDI (C-NODDI), or Standard Model Imaging (SMI) analyses, for one CN and one CI participant. Images are shown for the middle brain slice at three timepoints. **c** Results of the linear mixed-effects model of the association between whole-brain white matter ADI and time given by: 𝐴𝐷𝐼_𝑖𝑗_ ∼ β_0_ + β_𝐴𝑔𝑒_ × 𝐴𝑔𝑒_𝑖_ + β_𝑠𝑒𝑥_ × 𝑆𝑒𝑥_𝑖_ + β_𝑇𝑖𝑚𝑒_ × 𝑇𝑖𝑚𝑒_𝑖𝑗_ + β_𝐷𝑖𝑎𝑔𝑛𝑜𝑠𝑖𝑠_ × 𝐷𝑖𝑎𝑔𝑛𝑜𝑠𝑖𝑠_𝑖_ + β_𝑇𝑖𝑚𝑒×𝐷𝑖𝑎𝑔𝑛𝑜𝑠𝑖𝑠_ × 𝑇𝑖𝑚𝑒_𝑖𝑗_ × 𝐷𝑖𝑎𝑔𝑛𝑜𝑠𝑖𝑠_𝑖_ + 𝑏_𝑖_ + ϵ_𝑖𝑗_, Results are shown for each diagnosis group (i.e., CN and CI). CN and CI exhibit significant differences in axonal density/integrity as measured using ADI_NODDI_ or ADI_C-NODDI_. While the CN group maintained a relatively constant axonal density/integrity over time, the CI group exhibited decreases in ADI_NODDI_ and ADI_C-NODDI_, that is, decreased axonal density/integrity, over time. In contrast, ADI_SMI_ showed a slight increase over time. Full statistical results are shown in Supplementary Table S1.

Quantitatively, linear mixed-effects models were used to analyze longitudinal trajectories of whole-brain WM ADI values as a function of time from baseline. Model equations are provided in the corresponding figure legends. Each model included a Time × Diagnosis term to examine whether ADI trajectories differed between CN and CI groups, adjusting for relevant covariates (Fig. 2c). The results revealed that the CI group exhibited significantly lower ADI values compared to the CN group, with a larger effect size and stronger significance observed for C-NODDI [β_Diagnosis-CN_ = 0.63; *p* < 10^-6^] compared to NODDI [β_Diagnosis-CN_ = 0.44; *p* = 0.001], consistent with lower axonal density/integrity in CI. ADI_SMI_ did not show significant differences between groups [β_Diagnosis-CN_ = 0.22; *p* = 0.14]. Across all three dMRI models, time since baseline MRI was significantly and negatively associated with ADI values, demonstrating progressive reductions in axonal density/integrity over time. Importantly, the Time × Diagnosis interaction was significant and positively associated with both ADI_NODDI_ and ADI_C-NODDI_, indicating a steeper decline of axonal density in CI compared to CN (Fig. 2c). Similarly, the effect size and significance were notably greater for C-NODDI [β_Time_ = -0.21, *p* < 10^-8^; β_Time×Diagnosis-CN_ = 0.18, *p* < 10^-4^] compared to NODDI [β_Time_ = -0.18, *p* < 10^-9^; β_Time×Diagnosis-CN_ = 0.12, *p* = 0.001], highlighting the higher sensitivity of C-NODDI in differentiating ADI trajectories between CN and CI over time (Fig. 2c). No significant Time × Diagnosis interaction was observed for ADI_SMI_ [β_Time_ = 0.11, *p* = 0.046; β_Time×Diagnosis-CN_ = -0.07, *p* = 0.31]. Full regression results are provided in Supplementary Table S1.

### Association between baseline ADI and prospective changes in cognition

To investigate the association between baseline ADI and prospective changes in cognition, linear mixed-effects models were used, incorporating a three-way interaction term (Time × Diagnosis × ADI) and adjusting for relevant covariates (Fig. 3). Cognitive changes were assessed using the Mini-Mental State Examination (MMSE; Fig. 3a) and Clinical Dementia Rating–Sum of Boxes (CDR-SB; Fig. 3b). Thirteen participants were excluded from this analysis due to missing cognitive data (6 CN and 7 CI for MMSE; 5 CN and 8 CI for CDR-SB), either because they had no follow-up cognitive assessments or their first cognitive score was collected more than 0.1 years prior to baseline MRI (see Supplementary Fig. S2 and Fig. 3 a-b). The final cohort included 192 participants (111 CN and 81 CI for MMSE; 112 CN and 80 CI for CDR-SB). The average time difference between the first MMSE or CDR-SB measurement and the baseline MRI was 0.067 years (median: 0.023, SD: 0.167, range: –0.082 to 1.61) for MMSE and 0.068 years (median: 0.023, SD: 0.174, range: –0.082 to 1.74) for CDR-SB. The mean prospective follow-up duration was 1.69 years (median: 1.28, SD: 1.61, range: –0.058 to 5.04) for MMSE and 1.96 years (median: 1.99, SD: 1.60, range: –0.044 to 5.04) for CDR-SB. Analysis revealed that, as expected, the CN group maintained stable MMSE scores over time, while the CI group exhibited a progressive decline (Fig. 3c). Notably, this decline was steeper in participants with lower baseline ADI_C-NODDI_ (Fig. 3c, middle row, red prediction line). This association was statistically significant for C-NODDI [β_Time×ADI_ = 0.32, *p* = 0.03], but not for NODDI [β_Time×ADI_ = -0.06, *p* = 0.63] nor SMI [β_Time×ADI_ = 0.02, *p* = 0.85]. Similarly, while CDR-SB scores remained stable in the CN group, the CI group exhibited a prospective increase in CDR-SB scores, indicating increased dementia risk over time (Fig. 3d). However, the increase was steeper in participants with lower ADI_C-NODDI_ (Fig. 3d, middle row, red prediction line), whereas participants with higher ADI_C-NODDI_ values (green prediction line) maintained relatively stable CDR-SB scores despite a clinical diagnosis of cognitive impairment. Compared to NODDI [β_Time×ADI_ = -0.04, *p* = 0.60] and SMI [β_Time×ADI_ = 0.01, *p* = 0.86], C-NODDI demonstrated superior sensitivity in association with CDR-SB [β_Time×ADI_ = - 0.37, *p* < 10^-4^], supporting its potential as a prognostic biomarker for predicting dementia progression in individuals with cognitive impairment. Full results of the regression analyses are provided in Supplementary Table S2.

**Figure 3.**
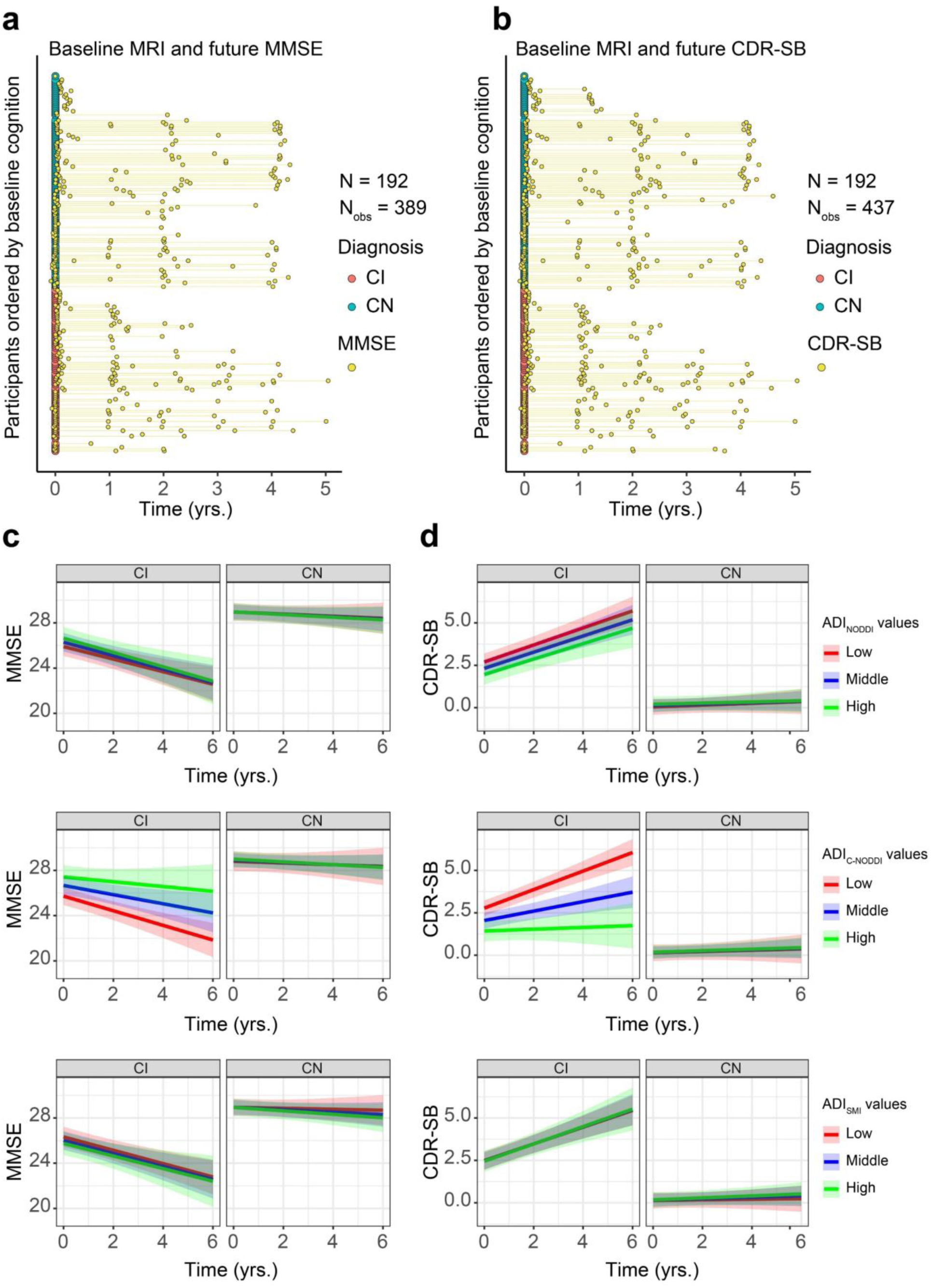
Baseline MRI measurements of axonal density/integrity, as measured using the axonal density index (ADI), predict prospective changes in cognition and dementia risk, as measured using the Mini-Mental State Examination (MMSE) and Clinical Dementia Rating-Sum of Boxes (CDR-SB) scores. Analyses were conducted using the following linear mixed-effects models: 𝑀𝑀𝑆𝐸/𝐶𝐷𝑅-𝑆𝐵_𝑖𝑗_ ∼ β_0_ + β_𝐴𝑔𝑒_ × 𝐴𝑔𝑒_𝑖_ + β_𝑠𝑒𝑥_ × 𝑆𝑒𝑥_𝑖_ + β_𝑇𝑖𝑚𝑒_ × 𝑇𝑖𝑚𝑒_𝑖𝑗_ + β_𝐷𝑖𝑎𝑔𝑛𝑜𝑠𝑖𝑠_ × 𝐷𝑖𝑎𝑔𝑛𝑜𝑠𝑖𝑠_𝑖_ + β_𝐴𝐷𝐼_ × 𝐴𝐷𝐼_𝑖_ + β_𝑇𝑖𝑚𝑒×𝐴𝐷𝐼_ × 𝑇𝑖𝑚𝑒_𝑖𝑗_ × 𝐴𝐷𝐼_𝑖_ + β_𝑇𝑖𝑚𝑒×𝐷𝑖𝑎𝑔𝑛𝑜𝑠𝑖𝑠_ × 𝑇𝑖𝑚𝑒_𝑖𝑗_ × ^𝐷𝑖𝑎𝑔𝑛𝑜𝑠𝑖𝑠^𝑖 ^+ β^𝐴𝐷𝐼×𝐷𝑖𝑎𝑔𝑛𝑜𝑠𝑖𝑠 ^× 𝐴𝐷𝐼^𝑖 ^× 𝐷𝑖𝑎𝑔𝑛𝑜𝑠𝑖𝑠^𝑖 ^+ β^𝑇𝑖𝑚𝑒×𝐷𝑖𝑎𝑔𝑛𝑜𝑠𝑖𝑠×𝐴𝐷𝐼 ^× 𝑇𝑖𝑚𝑒^𝑖𝑗 ^×^ 𝐷𝑖𝑎𝑔𝑛𝑜𝑠𝑖𝑠_𝑖_ × 𝐴𝐷𝐼_𝑖_ + 𝑏_𝑖_ + ϵ_𝑖𝑗_. (**a**, **b**) The longitudinal distributions of MMSE and CDR-SB anchored at the first diffusion MRI (dMRI) scan of each subject, with cognitively normal (CN) and cognitively impaired (CI) subjects color-coded. Both the CN and CI groups have similar longitudinal distributions, with many of the participants having over 4 years of cognitive measurement follow-up from baseline. **c** Predicted longitudinal changes in MMSE for CN and CI groups based on ADI_NODDI_, ADI_C-NODDI_ or ADI_SMI_ values. **d** Predicted longitudinal changes in CDR-SB for CN and CI groups based on ADI_NODDI_, ADI_C-NODDI_ or ADI_SMI_ values. While the CN group exhibits relatively constant MMSE and CDR-SB, the CI group shows significant decline in MMSE and increase in CDR-SB scores. Among ADI biomarkers, ADI_C-NODDI_ uniquely predicts differential trajectories within CI subjects, with higher baseline ADI_C-NODDI_ values predicting slower decline in cognition and lower increase in dementia risk (middle row, green prediction lines). These findings highlight the importance of maintaining axonal integrity, and the sensitivity of ADI_C-NODDI_ as an imaging biomarker for axonal integrity determination and prediction of cognitive decline and dementia risk. Full results are shown in Supplementary Table S2.

### Association between longitudinal changes in ADI and cognition

To investigate the association between longitudinal changes in ADI and changes in cognition and dementia risk, as measured using MMSE (Fig. 4a) or CDR-SB (Fig. 4b) scores, linear mixed-effects models were used, including a three-way interaction (Time × Diagnosis × ADI) and adjusting for relevant covariates. ADI values were centered at each subject’s baseline. Ten participants (5 CN and 5 CI) were excluded from the MMSE analysis and 11 participants (5 CN and 6 CI) were excluded from the CDR-SB analysis due to missing cognitive data. The final cohort included 195 participants (112 CN and 83 CI) for MMSE, with a mean follow-up interval of 0.955 years (median: 0; SD: 1.263; range: 0 to 4.167 years), and 194 participants (112 CN and 82 CI) for CDR-SB, with a mean follow-up interval of 0.935 years (median: 0; SD: 1.259; range: 0 to 4.167 years). Similar to the previous results (Fig. 2), the CN group exhibited higher ADI values compared to the CI group. Importantly, while the CN group showed minimal associations between longitudinal changes in ADI and changes in MMSE or CDR-SB scores, the CI group demonstrated that declines in ADI from baseline were associated with decreased MMSE scores and increased CDR-SB scores (Fig. 4c and d). This suggests that progressive axonal degeneration is associated with cognitive decline and increased dementia risk among individuals diagnosed with cognitive impairment. This association was statistically significant for ADI_NODDI_ [MMSE: β_ADI changes_ = 1.92, *p* < 10_-3_; β_Dignosis-CN×ADI changes_ = -2.05, *p* = 0.028; CDR-SB: β_ADI changes_ = -1.70, *p* < 10_-8_; β_Dignosis-CN×ADI changes_ = 1.67, *p* < 0.001] and ADI_C-NODDI_ [MMSE: β_ADI changes_ = 1.54, *p* = 0.005; β_Dignosis-CN×ADI changes_ = -1.98, *p* = 0.01; CDR-SB: β_ADI changes_ = -1.57; *p* < 10_-7_; β_Dignosis-CN×ADI changes_ = 1.52, *p* < 10^-4^]. These results showed that decreases in ADI were strongly linked to cognitive decline and dementia risk, with ADI_NODDI_ and ADI_C-NODDI_ outperforming ADI_SMI_. Full results of the regression analyses are presented in Supplementary Table S3.

**Figure 4.**
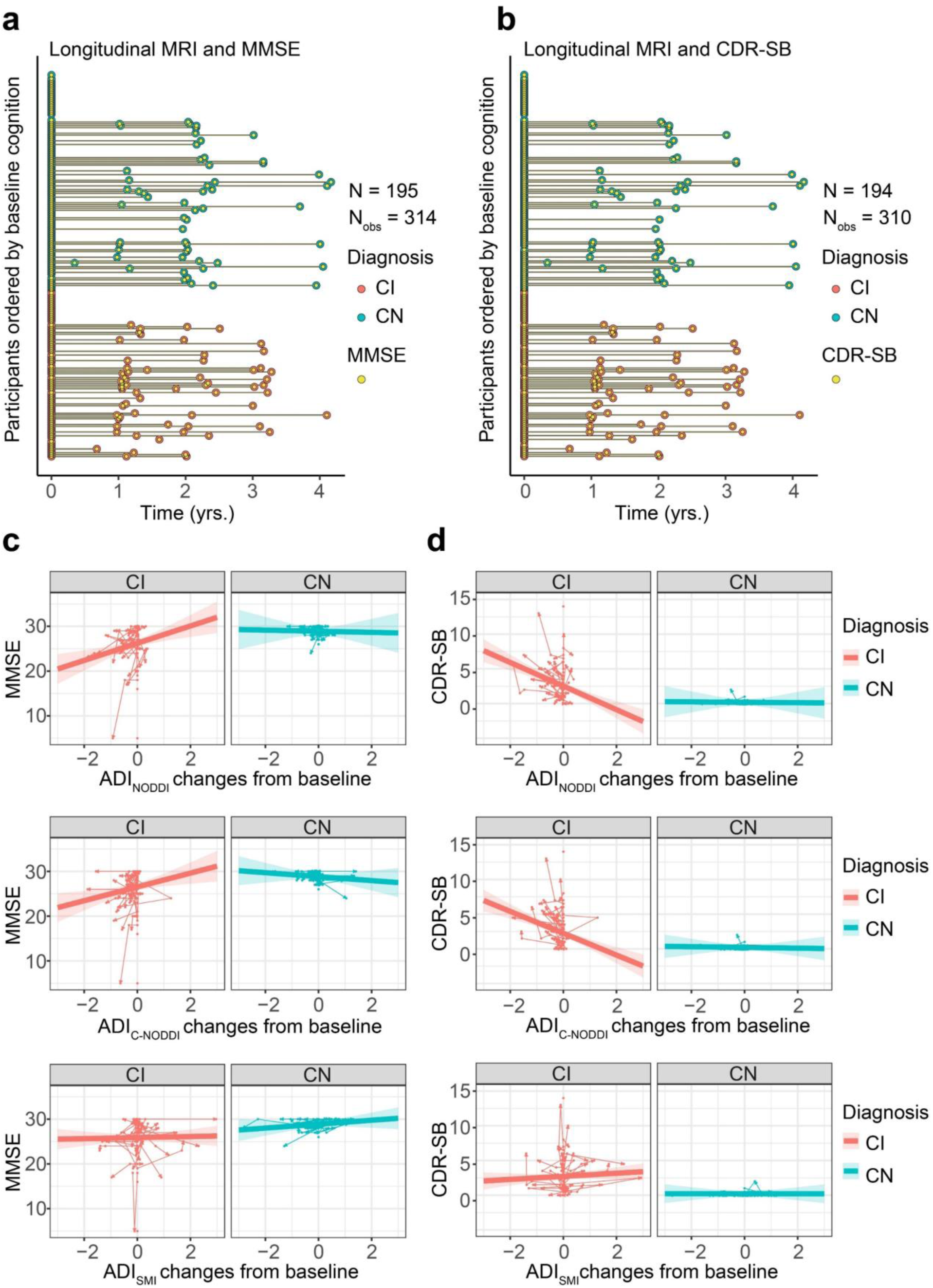
Changes in axonal integrity, as measured using the axonal density index (ADI), are associated with changes in cognition and dementia risk, as measured using the Mini-Mental State Examination (MMSE) and Clinical Dementia Rating-Sum of Boxes (CDR-SB) scores. Analyses were conducted using the following linear mixed-effects regression given by: 𝑀𝑀𝑆𝐸/𝐶𝐷𝑅- ^𝑆𝐵^𝑖𝑗 ^∼ β^0 ^+ β^𝑠𝑒𝑥 ^× 𝑆𝑒𝑥^𝑖 ^+ β^𝐴𝑔𝑒 ^× 𝐴𝑔𝑒^𝑖𝑗 ^+ β^𝐷𝑖𝑎𝑔𝑛𝑜𝑠𝑖𝑠 ^× 𝐷𝑖𝑎𝑔𝑛𝑜𝑠𝑖𝑠^𝑖 ^+ β^𝐴𝐷𝐼 𝑐ℎ𝑎𝑛𝑔𝑒𝑠 ^×^ 𝐴𝐷𝐼 𝑐ℎ𝑎𝑛𝑔𝑒𝑠_𝑖𝑗_ + β_𝐷𝑖𝑎𝑔𝑛𝑜𝑠𝑖𝑠×𝐴𝐷𝐼 𝑐ℎ𝑎𝑛𝑔𝑒𝑠_ × 𝐷𝑖𝑎𝑔𝑛𝑜𝑠𝑖𝑠_𝑖_ × 𝐴𝐷𝐼 𝑐ℎ𝑎𝑛𝑔𝑒𝑠_𝑖𝑗_ + β_𝐵𝑎𝑠𝑒𝑙𝑖𝑛𝑒 𝐴𝐷𝐼_ × 𝐵𝑎𝑠𝑒𝑙𝑖𝑛𝑒 𝐴𝐷𝐼_𝑖_ + β_𝐷𝑖𝑎𝑔𝑛𝑜𝑠𝑖𝑠×𝐵𝑎𝑠𝑒𝑙𝑖𝑛𝑒 𝐴𝐷𝐼_ × 𝐷𝑖𝑎𝑔𝑛𝑜𝑠𝑖𝑠_𝑖_ × 𝐵𝑎𝑠𝑒𝑙𝑖𝑛𝑒 𝐴𝐷𝐼_𝑖_ + 𝑏_𝑖_ + ϵ_𝑖𝑗_ To isolate within-subject effect, longitudinal ADI values are centered to the first ADI for each subject. (**a, b**) The longitudinal distribution of MMSE or CDR-SB scores in relation to the longitudinal MRI scans. To enable linear mixed-effects modeling, the MMSE and CDR-SB measurements were aligned to their closest diffusion MRI (dMRI) scans (see Supplementary Fig. S3 for original data distribution). **c** Individual trajectories show how changes in ADI are associated with changes in MMSE. Arrow directions indicate forward time progression. Decreases in ADI_NODDI_ and ADI_C-NODDI_ are associated with declines in MMSE for CI subjects, with ADI_C-NODDI_ and ADI_NODDI_ outperforming ADI_SMI_ in detecting such associations. **d** Individual trajectories show how changes in ADI are associated with changes in CDR-SB. Decreases in ADI_NODDI_ and ADI_C-NODDI_ are associated with increases in CDR-SB for CI subjects, with ADI_SMI_ failing to detect such association. Changes in ADI, that is, in axonal density/integrity are not associated with changes in MMSE or CDR-SB for CN subjects. However, it must be noted that the CN group exhibits both increase and decrease ADI from the baseline while the CI group predominately showed decreasing ADI over time. These findings underscore the implication of axonal integrity in cognition and dementia risk, and further emphasize the sensitivity of ADI_C-NODDI_ as an imaging biomarker for axonal integrity determination. Full results are shown in Supplementary Table S3.

### Comparison of ADI and CSF biomarkers of AD pathology performance

Subsets of participants with available multishell dMRI data and CSF biomarkers of AD pathology were used to compare the performance of ADI_C-NODDI_, Aβ_42/40_, tau and ptau_181_ in differentiating CN and CI trajectories and predicting cognitive decline. Panels a/d, b/e, and c/f in Fig. 5 directly compare the performance of CSF biomarkers and ADI_C-NODDI_ in differentiating CN and CI trajectories using matched subjects. As shown in Fig. 5, the CI group exhibited significantly lower ADI_C-NODDI_ values compared to the CN group, indicating lower axonal density in CI. Furthermore, ADI_C-NODDI_ trajectories revealed a steeper decline in axonal density in CI compared to CN. In contrast, while ptau_181_ [β_Diagnosis-CN_ = -0.24; *p* = 0.19] and tau [β_Diagnosis-CN_ = -0.22; *p* = 0.22] were elevated and Aβ_42/40_ [β_Diagnosis-CN_ = 0.39; *p* = 0.03] was reduced in CI compared to CN, none of these CSF biomarkers demonstrated clear differentiation in longitudinal trajectories between groups. Furthermore, baseline ADI_C-NODDI_ and Aβ_42/40_ provided comparable performance in predicting prospective cognitive decline and dementia risk among CI participants: individuals with lower baseline ADI_C-NODDI_ or Aβ_42/40_ values experienced greater cognitive decline and higher dementia risk (Fig. 6). Finally, while the CN group showed minimal association between longitudinal changes in ADI_C-NODDI_ and constant MMSE or CDR-SB scores, the CI group showed that decreased ADI_C-NODDI_ values are associated with decreased MMSE [β_ADI changes_ = 0.92; *p* = 0.11] or increased CDR-SB [β_ADI changes_ = - 1.16; *p* < 10^-3^] scores, indicating that axonal degeneration was associated decreased cognition and increased dementia risk in CI group (Fig. 7). The lack of statistical significance for MMSE likely reflects the smaller sample size in this subset compared to the analysis shown in Fig. 4. In contrast, the CI group showed that decreased Aβ_42/40_ values were unexpectedly associated with increased MMSE [β_Aβ42/40 changes_ = -4.1; *p* = 0.01] or decreaed CDR-SB [β_Aβ42/40 changes_ = 0.73; *p* = 0.56]. Full results of these regression analyses are presented in Supplementary Tables S4–S6.

**Figure 5.**
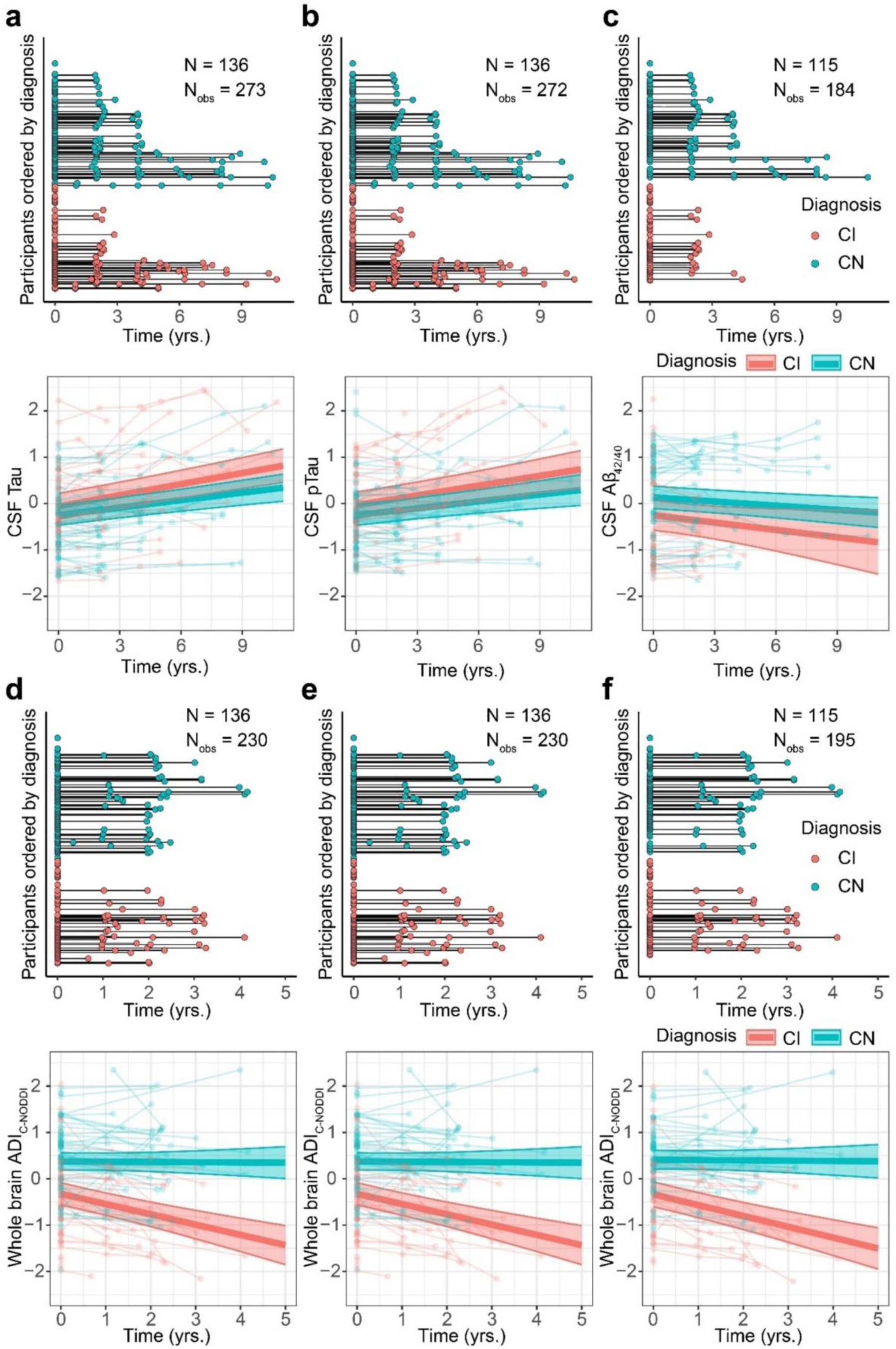
Comparison of CSF biomarkers and ADI_C-NODDI_ in differentiating longitudinal trajectories between the cognitively normal (CN) and cognitively impaired (CI) groups. Analyses were restricted to participants with both CSF biomarkers and diffusion MRI (dMRI). The comparison was restricted to ADI_C-NODDI_ as it is the best-performing dMRI biomarker in differentiating axonal degeneration trajectories between CN and CI in the previous analyses. **a**-**c** display the longitudinal data distributions for the three CSF biomarkers of AD pathology: tau, ptau_181_, and Aβ_42/40_. The trajectories of these CSF biomarkers show no significant differentiation over time between the CN and CI groups. **d**-**f** present the longitudinal dMRI data distribution for the ADI_C-NODDI_ biomarker from the same participants included in the above CSF analyses, for each CSF biomarker. In contrast to CSF biomarkers, the ADI_C-NODDI_ trajectories reveal significant differentiation between diagnosis groups, demonstrating the superior sensitivity of ADI_C-NODDI_ in detecting group differences over time compared to CSF biomarkers of AD pathology. Full results are shown in Supplementary Table S4.

**Figure 6.**
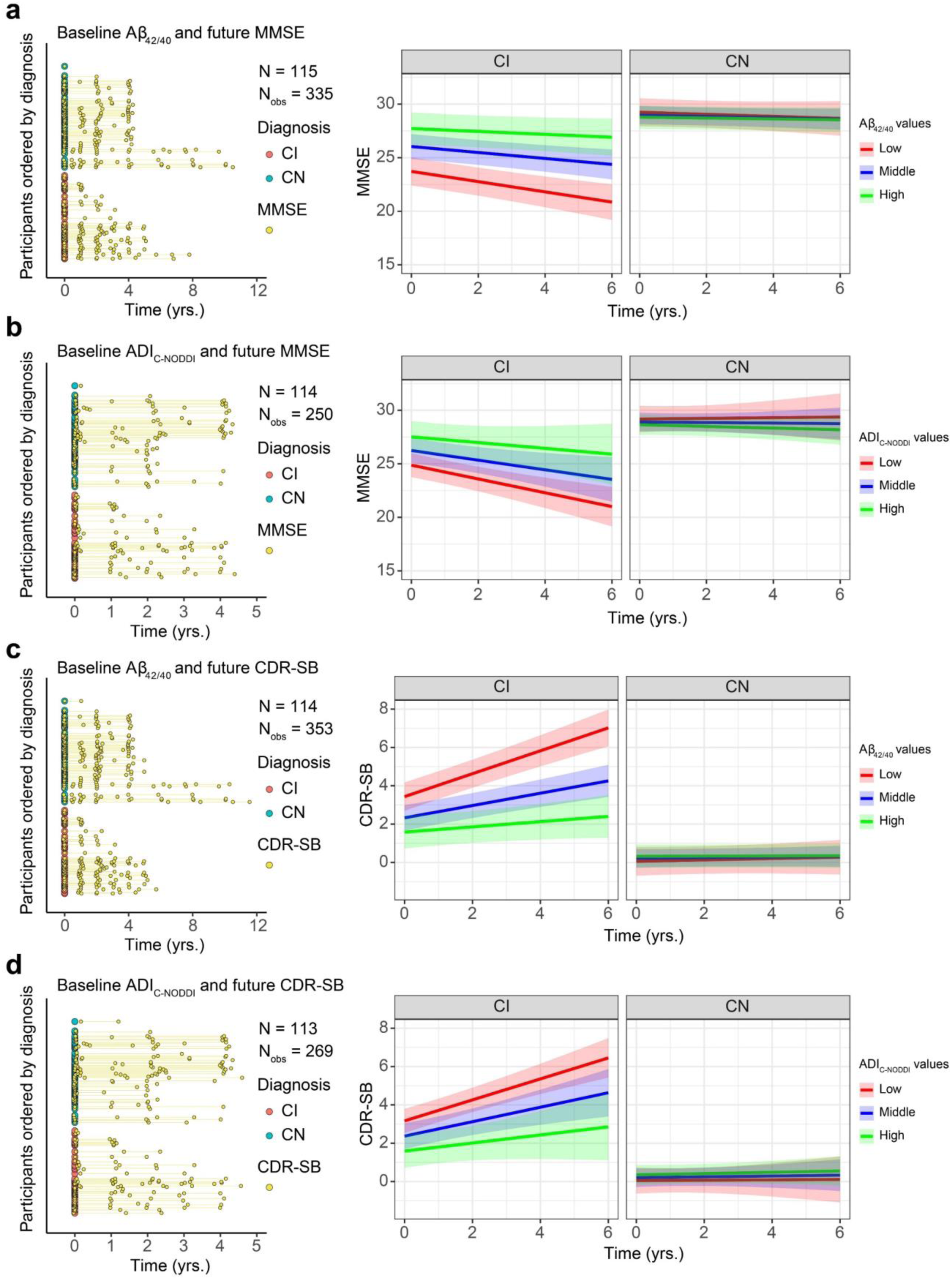
Comparison of baseline CSF Aβ_42/40_ and ADI_C-NODDI_ in predicting longitudinal changes in cognition and dementia risk in cognitively normal (CN) and cognitively impaired (CI) groups. Analysis was restricted to Aβ_42/40_ and ADI_C-NODDI,_ as they are the best performance from the previous analyses, and the same participants with available Aβ_42/40_ and ADI_C-NODDI_ biomarkers. The left panels show the longitudinal distributions of MMSE and CDR-SB anchored at the first dMRI scan of each subject, with CN and CI subjects color-coded. **a** displays the relationship between baseline Aβ_42/40_ levels and prospective MMSE scores, showing that a low Aβ_42/40_ ratio is significantly associated with faster MMSE decline in the CI group, but not in the CN group. **b** presents the relationship between baseline ADI_C-NODDI_ and future MMSE scores, where lower ADI_C-NODDI_ is also linked to faster MMSE decline in the CI group, though the association is close to significance (p = 0.07). **c** shows the relationship between baseline Aβ_42/40_ and prospective CDR-SB scores, with lower Aβ_42/40_ ratios significantly predicting a faster increase in CDR-SB in the CI group, but not in the CN group. **d** demonstrates that lower baseline ADI_C-NODDI_ is significantly associated with a faster increase in CDR-SB in the CI group, but not in the CN group. This analysis shows that both the Aβ_42/40_ and ADI_C-NODDI_ biomarkers exhibit similar performances in predicting prospective cognitive decline and dementia risk. Full results are shown in Supplementary Table S5.

**Figure 7.**
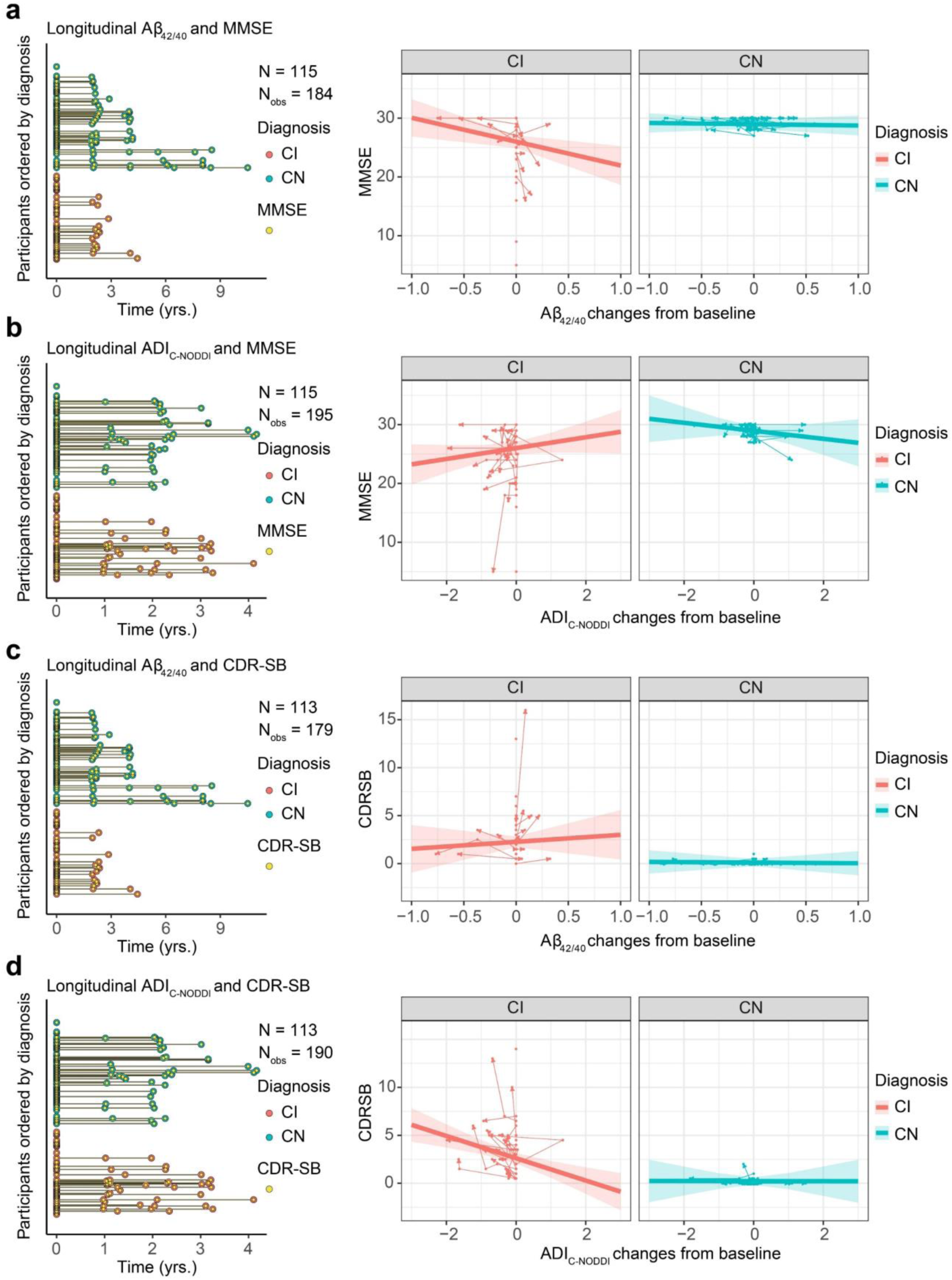
Comparison of changes in Aβ_42/40_ and ADI_C-NODDI_ changes in predicting longitudinal MMSE and CDR-SB changes. **a** The longitudinal distribution of MMSE scores aligned with the nearest Aβ42/40 measurements for linear mixed-effects modeling. Decreases in Aβ_42/40_ are significantly associated with increased MMSE in CI subjects but not in CN subjects. **b** The longitudinal distribution of MMSE scores aligned with corresponding ADI_C-NODDI_ for the same subjects in panel **a**. In comparison, decreases in ADI_C-NODDI_ are associated with MMSE decline in CI subjects (close to significant) but not in CN subjects. **c** Decreases in Aβ_42/40_ are associated with decreasing CDR-SB in CI subjects but not in CN subjects. **d** Decreases in ADI_C-NODDI_ are significantly associated with CDR-SB increase in CI subjects but not in CN subjects. The counter-intuitive results for Aβ_42/40_ in **a** and **c** are likely due to the small sample size, while the results for ADI_C-NODDI_ in **b** and **d** are similar to Fig. 4. Full results are shown in Supplementary Table S6.

As a sensitivity analysis, we repeated the analyses in Fig. 2-4 after excluding the 14 participants with a clinical diagnosis of AD from the CI group, therefore restricting the CI group to MCI participants only. Conclusions remained relatively unchanged and can be found in Supplementary Tables S7–S9 and Figures S4-S6. Additionally, while group differences in baseline ADI_C-NODDI_ were significant between CN and MCI (p < 10^-4^), the differences in all CSF biomarkers between the groups were not significant (*p* > 0.08).

## Discussion

### Summary of main findings

To our knowledge, this is the first study leveraging longitudinal multishell dMRI data to examine the implication of axonal degeneration in normative aging, MCI, or AD. By integrating dMRI, CSF biomarkers, and cognitive assessments from the ADNI study, we demonstrate that: *i*) axonal density/integrity, as measured using ADI, particularly when measured using the recently proposed C-NODDI model, declines more steeply in CI individuals compared to CN controls. *ii*) These changes in ADI are predictive of future cognitive decline and dementia risk. *iii*) C-NODDI predictivity outperforms other dMRI models (NODDI and SMI) and provides predictive power that is comparable to or better than CSF biomarkers, with the added benefit of being noninvasive and spatially specific.

### Axonal degeneration as a potential core feature of sporadic Alzheimer’s disease

Our results provide evidence supporting the role of axonal degeneration in the progression of cognitive impairment (in MCI and AD) and its association with cognitive decline. Postmortem histological studies indicate that WM degeneration, including axonal loss, is a primary consequence of aging and AD, associated with cognitive decline, motor impairments, and neurodegenerative disorders (39). These alterations in WM microstructure have been observed in early-onset autosomal-dominantly inherited AD occurring years before the symptom onset (9). Such WM alterations are believed to be associated with primary AD pathology and microglia activity in the brain (20). By probing one specific component of WM, our work revealed lower axonal density in patients with cognitive impairment, in agreement with previous cross-sectional-based work that used NODDI (40–43). Moreover, axonal density, as measured using our ADI imaging biomarker, predicted future cognitive decline and dementia risk, and longitudinal changes in ADI were also associated with concurrent changes in cognitive performance. These results provide new evidence that progressive axonal degeneration is likely a key driver of cognitive deterioration in the early stages of AD and highlight the potential of ADI as a sensitive, noninvasive biomarker for tracking disease progression.

Axonal degeneration is a complex, multi-step process that is primarily driven by disruptions in cellular homeostasis, energy deficits, and altered intracellular signaling pathways (44–46). At the molecular level, it involves the breakdown of key components of the axon, such as microtubules and neurofilaments, which are crucial for maintaining structural integrity and efficient axonal transport. Impaired axonal transport, often due to mitochondrial dysfunction or the accumulation of toxic proteins like tau, leads to the accumulation of waste products and energy depletion, further exacerbating degeneration (47–49). Additionally, the activation of signaling pathways, including those mediated by calpains and other proteases, can result in the cleavage of structural proteins and the destabilization of the axonal cytoskeleton, accelerating the process of axonal disintegration (50, 51). These molecular events, often triggered by neuroinflammation and oxidative stress, contribute to the progressive loss of axonal function and ultimately, neuronal death, brain atrophy and concomitant cognitive and motor dysfunctions. Our results indicate that patients who have a clinical diagnosis of cognitive impairment, but greater axonal density, are spared from accelerated cognitive decline and dementia risk. These original results underscore the importance of axonal health and call for further investigations into axonal degeneration as a potential core feature of AD.

### Clinical implications and significance

The findings of this study have important implications for both clinical research and our broader understanding of AD pathophysiology. The ability to detect axonal degeneration, measured through ADI, prior to significant cognitive decline or overt brain atrophy underscores its potential as a critical early marker of disease progression. This is particularly relevant for identifying individuals in the prodromal or pre-clinical stages of AD, when therapeutic interventions are more likely to be effective. One of the most compelling findings is that ADI effectively differentiated longitudinal trajectories between CN and CI individuals, establishing its potential as a monitoring biomarker. This capability will allow clinicians to track disease progression dynamically and identify patients who are undergoing steeper axonal density decline, enabling more timely interventions. Furthermore, our observation that some CI individuals exhibited relatively high ADI levels and slower cognitive decline suggests that axonal density may serve as an indicator of cognitive resilience or reserve. These findings support the use of ADI not only for disease monitoring and early diagnosis but also for disease phenotyping and patient stratification. The degree of axonal degeneration could help identify subgroups with distinct clinical trajectories and those more likely to respond to targeted therapies. This has important implications for clinical trial design, as it may improve inclusion criteria, reduce heterogeneity, and increase the likelihood of detecting treatment effects. In terms of intervention, our results reinforce the importance of developing treatments specifically aimed at preserving or restoring axonal integrity. Whether through pharmacological agents, lifestyle modifications (*e.g.*, physical activity, diet), or cognitive training, interventions that enhance or maintain axonal health could delay or mitigate the progression of cognitive symptoms. ADI offers a quantifiable imaging biomarker to monitor the effectiveness of these strategies over time, which could accelerate the development and validation of WM-targeted therapies. Finally, the observation that CN and MCI participants had similar CSF biomarker profiles, but significantly different ADI levels highlight the added value of dMRI-based markers. Higher axonal integrity in the presence of AD pathology may reflect a protective factor, contributing to delayed symptom onset. This supports the notion that ADI could serve as a marker of cognitive reserve and may ultimately guide the development of more personalized, stage-specific treatment strategies. Our results underscore the utility of ADI, particularly when derived using C-NODDI, as a sensitive and clinically meaningful biomarker for early detection, disease monitoring, patient stratification, and therapeutic targeting in AD.

### Comparison to CSF biomarkers of AD pathology

Another major finding of our investigation is that our biomarker of axonal density, ADI_C-NODDI_, provided similar or superior results to CSF biomarkers in differentiating CN from CI, and predicting cognitive decline and dementia risk. MRI allows for direct assessment of microstructural changes in the brain, including myelin integrity and axonal loss, which can occur early in the disease and reflect ongoing neurodegeneration (52–55). In contrast, CSF biomarkers, while informative, represent biochemical changes that may occur downstream of initial neurodegenerative events and may not fully capture the complexity of neurodegenerative processes, particularly in the early stages of AD when axonal degeneration may precede significant changes in CSF composition. Although not the focus of current work, dMRI provides high spatial resolution, enabling the detection of topographical patterns of axonal density across different brain regions. These regional patterns are often differentially linked to cognitive decline and disease progression, offering more granular information than CSF biomarkers which reflect a global concentration of proteins. The ability to capture this detailed, localized information in dMRI allows for more precise differentiation between healthy controls and individuals with early AD pathology, enhancing its predictive power for cognitive decline and dementia risk, as shown in our and others’ recent work (56, 57). Furthermore, dMRI is noninvasive and easily repeatable, allowing for longitudinal monitoring of disease progression without the need for invasive procedures, like CSF collection through lumbar puncture. The noninvasive nature of dMRI makes it a more patient-friendly and accessible option for regular assessment, posing particular advantages in clinical settings where repeated testing is necessary. While PET imaging also provides topographical insights into brain activity and pathology, it remains costly, invasive, and less accessible. In addition, PET imaging suffers from low resolution, making detection of subtle microstructural or chemical changes, including in small brain regions, challenging. Finally, while there are growing efforts to develop plasma biomarkers for AD, which offer the advantage of accessibility and scalability (58–62), they still require further validation and cannot provide the same anatomical specificity as imaging-based approaches. In this context, ADI_C-NODDI_ could serve as a complementary biomarker, offering spatially resolved, mechanistically informative insights into neurodegeneration. Further work is needed to validate ADI in multi-cohort data with diverse populations and to explore its integration with fluid biomarkers to enhance early diagnosis, disease staging and phenotyping, and treatment monitoring in AD (33, 40, 56, 57).

### Improving the AT(N) framework

The AT(N) framework, which categorizes AD biomarkers into amyloid (A), tau (T), and neurodegeneration (N), has proven instrumental in advancing our understanding of the disease’s pathophysiology (6). However, recent studies have begun to emphasize the role of WM integrity, particularly axonal degeneration, as an important factor in the neurodegenerative process (11). While the N component of the AT(N) framework currently focuses on markers such as hippocampal atrophy and CSF neurofilament light chain, our work suggests that axonal degeneration may play a pivotal role in early disease stages, before significant cortical atrophy becomes evident. Notably, alterations in WM integrity, as measured through established MRI techniques, have been associated with both amyloid and tau pathology (55, 63, 64), providing a potential bridge between the pathological hallmarks of AD and the clinical manifestation of cognitive impairment. Incorporating axonal degeneration, as measured using C-NODDI, into the AT(N) framework could offer new insights into the timing and progression of neurodegeneration, potentially enhancing early detection and therapeutic intervention strategies. Furthermore, combined with protein biomarkers, this imaging biomarker has the potential to improve the AT(N) framework in accurate prediction of disease progression, AD diagnosis, and patient stratification in clinical trials (35, 40, 65).

### Strengths and limitations

Changes in axonal density were markedly depicted using our C-NODDI dMRI method as compared to other state-of-the-art techniques, including NODDI and SMI. However, while compelling, these biophysical tissue models involve a high-dimensional inverse problem, making derived parameters, especially ADI, dramatically sensitive to the impact of experimental conditions and system noise. This impedes their sensitivity to early changes due to pathology, as clearly seen in this work. However, C-NOODI provides physiologically realistic ADI_C-NODDI_ values that exhibit a stronger correlation with the neurofilament light chain (NfL), a plasma biomarker of axonal degeneration (38), with, as expected, lower ADI_C-NODDI_ values corresponded to higher NfL levels. This technical advancement and our current results underscore the utility of ADI_C-NODDI_ as a powerful, noninvasive, and cost-effective biomarker to help elucidate the mechanisms of axonal degeneration, early diagnosis, and monitoring cognitive decline due to MCI and AD.

While our study offers promising and novel insights into the role of axonal degeneration in the AD spectrum and demonstrates the utility of our C-NODDI dMRI method in detecting early axonal changes and their implication in cognitive decline, there are limitations to consider. First, although our results show that C-NODDI-derived ADI performs similarly or superiorly to CSF biomarkers, dMRI still relies on model assumptions that may limit its accuracy in certain populations, particularly in those with advanced disease or comorbid conditions. The sensitivity of our method to detect subtle changes in axonal integrity may also be influenced by the quality of the MRI data, as multishell diffusion imaging is prone to noise and artifacts, which could potentially affect the interpretation of results. Therefore, sophisticated analyses and expertise, as used here, are required to mitigate these potential issues. Further, we only considered whole brain WM regions in our analysis. However, C-NODDI offers valuable spatial information that can allow for the examination of the spatial distribution of axonal degeneration, even subtle changes, across different regions of the WM, helping to capture early signs of degeneration. Nevertheless, despite focusing on whole brain WM, C-NODDI was able to depict global variations in axonal integrity, enhancing our understanding of how these changes may be linked to cognitive decline and dementia risk. Studies focusing on the spatial pattern and involving other cognitive assessments are warranted in the future. Additionally, while our study focuses on longitudinal changes, the relatively short duration of the follow-up period may not fully capture the long-term trajectory of axonal degeneration and its relationship with cognitive decline, particularly in the preclinical stage of AD. With the continuing progress of the ADNI study, we hope to enhance our results when future visits become available. Furthermore, the generalizability of our findings may be limited by the sample size and the specific cohort used, which mainly includes participants from the ADNI database, and may not fully represent the broader spectrum of AD and other neurodegenerative diseases. Moreover, males had a longer follow-up than females, therefore a bias toward male participants may influenced the overall results of this study. Lastly, while dMRI offers a noninvasive and repeatable means of tracking disease progression, it remains resource-intensive compared to CSF biomarkers, requiring specialized equipment and expertise. However, we remain hopeful about the expanding accessibility of dMRI to the general population with the technical advancement in low-field portable MRI. Despite these limitations, our original findings highlight the implication of axonal degeneration in AD and the potential of C-NODDI to provide valuable insights into the early stages of AD, underscoring the need for further validation in larger, more diverse cohorts with extended follow-up periods.

## Methods

### Participants

Participants were drawn from the ADNI database (https://adni.loni.usc.edu/). The ADNI initiative was started in 2003 under Michael W. Weiner, MD, and was a private-public partnership funded by private companies as well as the National Institutes of Health (NIH) and National Institute on Aging (NIA) for the purpose of developing clinical, imaging, genetic, and biochemical biomarkers for early detection of AD. Inclusion criteria were participants who had multi-shell diffusion scans on Siemens scanners. Baseline and longitudinal dMRI scans were obtained across all available participants. A subset of these participants also had CSF fluid biomarkers. All participants signed consent forms, and the study design was approved by the IRB of data-collection institutions. For more information, see www.adni-info.org.

### dMRI imaging and processing

Each participant underwent whole brain dMRI scans using a 3 T Siemens scanner. MAGNETOM Skyra, MAGNETOM Prisma, and MAGNETOM Prisma^fit^ were used depending on the scanning site. Multi-shell diffusion MRI data were collected with a repetition time of 3400 ms and an echo time of 71 ms. Each scan included 127 separate diffusion-weighted images: 13 with *b* = 0 s/mm², 6 with *b* = 500 s/mm², 48 with *b* = 1000 s/mm², and 60 with *b* = 2000 s/mm². The dMRI scans were pre-processed. First, the raw imaging dataset was processed in Python to convert individual DICOM images into 4-D NIfTI files and standardized in format. Images were subsequently denoised using the MP-PCA MATLAB toolbox. Brain extraction and linear registration of each diffusion-weighted image to the b_0_ image were performed using the FMRIB Software Library (FSL) (66). Eddy current-induced distortions and participant movements were corrected using FSL, and ADI maps were then derived using the NODDI, C-NODDI, or SMI models (36–38). Lastly, derived parameter maps were registered to the JHU-ICBM-T_2_-2mm template from the JHU DTI atlases using linear and nonlinear FSL FLIRT and FNIRT functions. White matter voxels were defined based on a threshold of a WM mask with a probability higher than 95%. The whole brain WM ADI values were then calculated.

### CSF biomarkers

Lumbar CSF samples were collected and stored at −80°C at the ADNI Biomarker Core at the University of Pennsylvania School of Medicine. The established CSF biomarkers of amyloid-βeta ratio (Aβ_42/40_), total tau protein (tau), and phosphorylated-tau (pTau_181_) were obtained from a subset of ADNI participants who had multishell dMRI scans. All CSF biomarker values were obtained from the *UPENN CSF Biomarkers Roche Elecsys [ADNI1,GO,2,3]* CSV file and detailed assay methods can be found in the ADNI publication *UPENN CSF Biomarkers Roche Elecsys Methods [ADNI1,GO,2,3]* file. Further collection details can be found through ADNI Documentation (https://adni.loni.usc.edu/help-faqs/adni-documentation/).

### Cognitive scores

The Mini-Mental State Examination (MMSE) and Clinical Dementia Rating - Sum of Boxes (CDR-SB) scores were used in this study to assess global cognition and dementia risk over time. MMSE and CDR-SB values were drawn from the *ADNIMERGE - Key ADNI tables merged into one table [ADNI1, GO, 2, 3]* file. Further information on the cognitive assessment can be found on the ADNI site.

### Statistical analysis

All statistical analyses were conducted in RStudio 4.3. The longitudinal multishell dMRI, CSF, and cognitive data and linear mixed-effects regression analyses were used for the following objectives: (*i*) to examine longitudinal changes in axonal density and integrity, as measured by ADI_NODDI_, ADI_C-NODDI_, or ADI_SMI_; (*ii*) to investigate whether baseline ADI predicts prospective changes in cognition, as measured by MMSE and CDR-SB scores; and (*iii*) to assess the association between longitudinal changes in ADI and longitudinal changes in MMSE and CDR-SB scores. Similar analyses were conducted using CSF biomarkers of AD pathology (Aβ_42/40_, tau, ptau_181_), which were available for a subset of participants with both multishell dMRI data and CSF biomarkers. To ensure a fair comparison, the results were compared to ADI_C-NODDI_ using the closest subset of full multishell dMRI data. For simplicity, the comparison was restricted to CSF Aβ_42/40_ and ADI_C-NODDI_, as these biomarkers showed superior performance compared to the other CSF and dMRI biomarkers in the previous analyses. All analyses included interaction terms with diagnosis groups (CN, CI) to delineate the group difference, adjusting for relevant covariates, namely age and sex. The regression models are displayed in the legends of the corresponding figures.

To verify that our results were not entirely driven by the inclusion of the AD patients, all the abovementioned analyses were repeated by excluding the AD patients from the CI group. In addition, to investigate whether axonal integrity confers cognitive reserve, baseline differences between the CN and MCI groups were compared for each CSF biomarker of AD pathology and ADI_C-NODDI_, accounting for age and sex as relevant covariates.

### Study approval

All participants signed consent forms, and the study design was approved by the IRB of the National Institute on Aging. For more information, see www.adni-info.org.

### Data availability

Tubular Data for all data points in graphs and analysis codes are available from GitHub (https://github.com/mrpadunit).

## Author contributions

ZG designed the study, performed experiments, ran analyses, and wrote and edited the manuscript. JL and AG performed experiments and edited the manuscript. JB, NF, AdR, NZ, and AT edited the manuscript. RdC, JE, LF provided intellectual discussion and edited the manuscript. MB conceived the study, designed experiments, and wrote and edited the manuscript. All authors provided comment on the final manuscript.

## Acknowledgments

Data collection and sharing for this project was funded by the Alzheimer’s Disease Neuroimaging Initiative (ADNI) (National Institutes of Health Grant U01 AG024904) and DOD ADNI (Department of Defense award number W81XWH-12-2-0012). ADNI is funded by the National Institute on Aging, the National Institute of Biomedical Imaging and Bioengineering, and through generous contributions from the following: AbbVie, Alzheimer’s Association; Alzheimer’s Drug Discovery Foundation; Araclon Biotech; BioClinica, Inc.; Biogen; Bristol-Myers Squibb Company; CereSpir, Inc.; Cogstate; Eisai Inc.; Elan Pharmaceuticals, Inc.; Eli Lilly and Company; EuroImmun; F. Hoffmann-La Roche Ltd and its affiliated company Genentech, Inc.; Fujirebio; GE Healthcare; IXICO Ltd.; Janssen Alzheimer Immunotherapy Research & Development, LLC.; Johnson & Johnson Pharmaceutical Research & Development LLC.; Lumosity; Lundbeck; Merck & Co., Inc.; Meso Scale Diagnostics, LLC.; NeuroRx Research; Neurotrack Technologies; Novartis Pharmaceuticals Corporation; Pfizer Inc.; Piramal Imaging; Servier; Takeda Pharmaceutical Company; and Transition Therapeutics. The Canadian Institutes of Health Research is providing funds to support ADNI clinical sites in Canada. Private sector contributions are facilitated by the Foundation for the National Institutes of Health (www.fnih.org). The grantee organization is the Northern California Institute for Research and Education, and the study is coordinated by the Alzheimer’s Therapeutic Research Institute at the University of Southern California. ADNI data are disseminated by the Laboratory for Neuro Imaging at the University of Southern California. This work was supported by the National Insititute on Aging Intramural Research program.

